# Reverse-Engineering the Benefits of Stereotypies in Autism: Vibrating Vest Design

**DOI:** 10.1101/2025.03.05.25323041

**Authors:** Gabriela Franco, Peter Lin, Emily Liu, Selim Uzgoren, Abhinav Vetcha, Jonathan T. Pierce, Audrey C. Brumback

**Affiliations:** Department of Biomedical Engineering, The University of Texas at Austin, Austin, Texas USA; Department of Neuroscienc, The University of Texas at Austin, Austin, Texas USA; Center for Learning and Memory, The University of Texas at Austin, Austin, Texas USA; Departments of Neurology and Pediatrics at Dell Medical School, The University of Texas at Austin, Austin, Texas USA

**Keywords:** autism spectrum disorder, stereotypies, wearables, vibrotactile, intellectual disability

## Abstract

**Objective:** Individuals with autism may perform repetitive or stereotyped rhythmic movements known as “stereotypies” or “stims.” Per first-person reports, stereotypies provide benefits including sensory and emotional regulation. These movements are not often convenient to engage in and for some may be self-injurious. We propose the design of a device that discreetly provides rhythmic stimulation through vibrating motors that are incorporated into a vest designed to be worn under clothing.

**Methods:** We designed a vest that provides sensory stimulation through vibrations from 18 coin motors arranged on the interior surface of a compression vest. The vibration frequency and duty cycle are controlled by the user via a Bluetooth smartphone application, allowing them to optimize the stimulation for their unique needs. The vest is designed to be worn discreetly under the user’s clothing. It is rechargeable and can be used at full power for up to 4 hours. We performed experiments to quantify target values for the vibration pressure, signal frequency, signal duty cycle, battery life, noise level, and temperature.

**Results:** Our device accurately controls frequency and duty cycle with marginal error and meets all our engineering requirements except for battery life. It also conforms to ethical and regulatory guidelines.

**Conclusion:** Future work, such as variable device weight, pressure and temperature control, and vibration patterns synchronized to music, could refine the product and deliver more value to the user. Significance: We have designed a vest using vibration to provide rhythmic sensory input to supplement the benefits of stereotyped movements.

## INTRODUCTION

Autism spectrum disorder (ASD) is a neurodevelopmental condition characterized by the co-occurrence of (1) atypical social communication and (2) restrictive and repetitive interests and behaviors [1]. ASD affects over 1 in 40 children in the United States [2]. There are no extant medical treatments for the core symptoms of ASD. Stereotyped movements are one prevalent type of repetitive behavior in ASD [3]. These “motor stereotypies,” colloquially referred to as “stimming,” are rhythmic and repetitive movements, often with a fixed pattern and constant frequency. Examples include rocking, flapping, finger-flicking, spinning, and jumping [4–6]. Almost half of children with ASD have at least one type of stereotypy [7,8].

The presence of motor stereotypies may be associated with differences in the function of cortico-striatal-thalamo-cortical pathways, which are central to cognitive control [9,10]. Psychiatric conditions involving cognitive dysfunction and emotional dysregulation also exhibit dysfunction in this pathway [6,11]. People who exhibit stereotypies often report that making the repetitive movements improves focus and sensory processing and decreases anxiety [12]. How repetitive movements and the resulting rhythmic sensory signals improve brain function is unknown. Intriguingly, a common finding in people with ASD is dysregulated brain rhythms [13]. Well-timed brain rhythms are necessary for effective sensory processing and attention [14,15]. Thus, one possibility for how performing stereotyped movements might benefit the person is that the rhythmic motor signal and/or the rhythmic sensory feedback produced may entrain brain rhythms and thereby improve focus and sensory processing (Appendix A) [12].

The positive aspects of motor stereotypies are tempered by the negatives [16]. First, it is not often convenient to perform rhythmic sterotyped movements at the time they might be most needed (e.g., sitting in class, riding the bus, etc.). In many societies, stereotypies are unfortunately socially stigmatized and can reduce involvement within the community. Stereotypies can also take the form of self-injurious behaviors, such as hand-banging, skin picking, self-scratching, and self-biting [17–19].

Therefore, identifying mechanisms to provide the benefit of the stereotyped movement without the need to perform the movements is critical for harnessing their benefit as a therapy. If the rhythmic sensory signal caused by the movements is part of the beneficial mechanism, we hypothesized that providing rhythmic sensory inputs exogenously could produce the benefit of stereotypies without the need for the movements themselves. To test this hypothesis in future studies, we developed a vibrating compression vest designed to be worn under clothing.

### Current Solutions and Treatment Options

Weighted compression vests and inflatable deep pressure vests have been used to provide a sense of safety and comfort [20,21]. There are various other weighted products available such as blankets, backpacks, lap pads, and toys. Massage pillows, sensory pads, vibrating toys, and other vibration therapies are described as calming for individuals with ASD [22–24]. Full-body vibration and vibroacoustic music may decrease the frequency of some stereotypies in people with autism [25,26]. Besides being inconvenient, there may be health drawbacks to using whole-body vibration therapy for an extended period [27]. Transcutaneous vagus nerve stimulation (tVNS), a treatment for epilepsy and other neuropsychiatric conditions, has been proposed as a potential therapy for ASD [28]. This is grounded in differences in autonomic nervous system function in people with autism and other neurodevelopmental conditions [29–31]. Adverse effects such as hoarseness and cough are common with VNS, which may limit its use [32].

### Analyses

#### Market, Stakeholder, and Customer Needs Analyses

Of the approximately 3% of the population with autism, approximately half exhibit motor stereotypies [8]. Assuming a 1% market penetration, we extrapolate 100,000 possible users. We expect this device’s beneficiaries to be primarily individuals who have motor stereotypies and would like to have an additional source of stimulation for comfort and focus.

#### Need Statement

This vest addresses disruptive motor stereotypies in people with autism by potentially providing the neurological benefits of stimming without the need to generate repetitive movements (Tables I – III).

**Table I.** General Need Criteria.

| Need | Explanation |
| --- | --- |
| Efficacious | The device should function well as a substitute for a motor stereotypy. |
| Discreet | The device should be less obvious than a motor stereotypy. |
| Easy to use | The device should be simple and intuitive to use. |
| Durable | The device should be resistant to adverse environmental conditions. |
| Portable | The device should be easily transported with the user and integrated into daily life. |
| Affordable | The device should be affordable so that it is accessible to more people. |
| Comfortable | The device should feel comfortable for long-term use. |

**Table II.** Specific Need Criteria.

| Must-Haves | Nice-to-Haves |
| --- | --- |
| Efficacious at mimicking neurological feedback | Provides deep pressure for calming sensation |
| Comfortable and nonstimulatory until powered on | Covers a large surface area for better stimulation |
| Safe to wear | Long battery life for reliable daily use |
| Easy to use and intuitive to operate | Durable to withstand various conditions during use |
| Discreet enough to be nondistracting to user and others | Adjustable size for user comfort |

**Table III.**
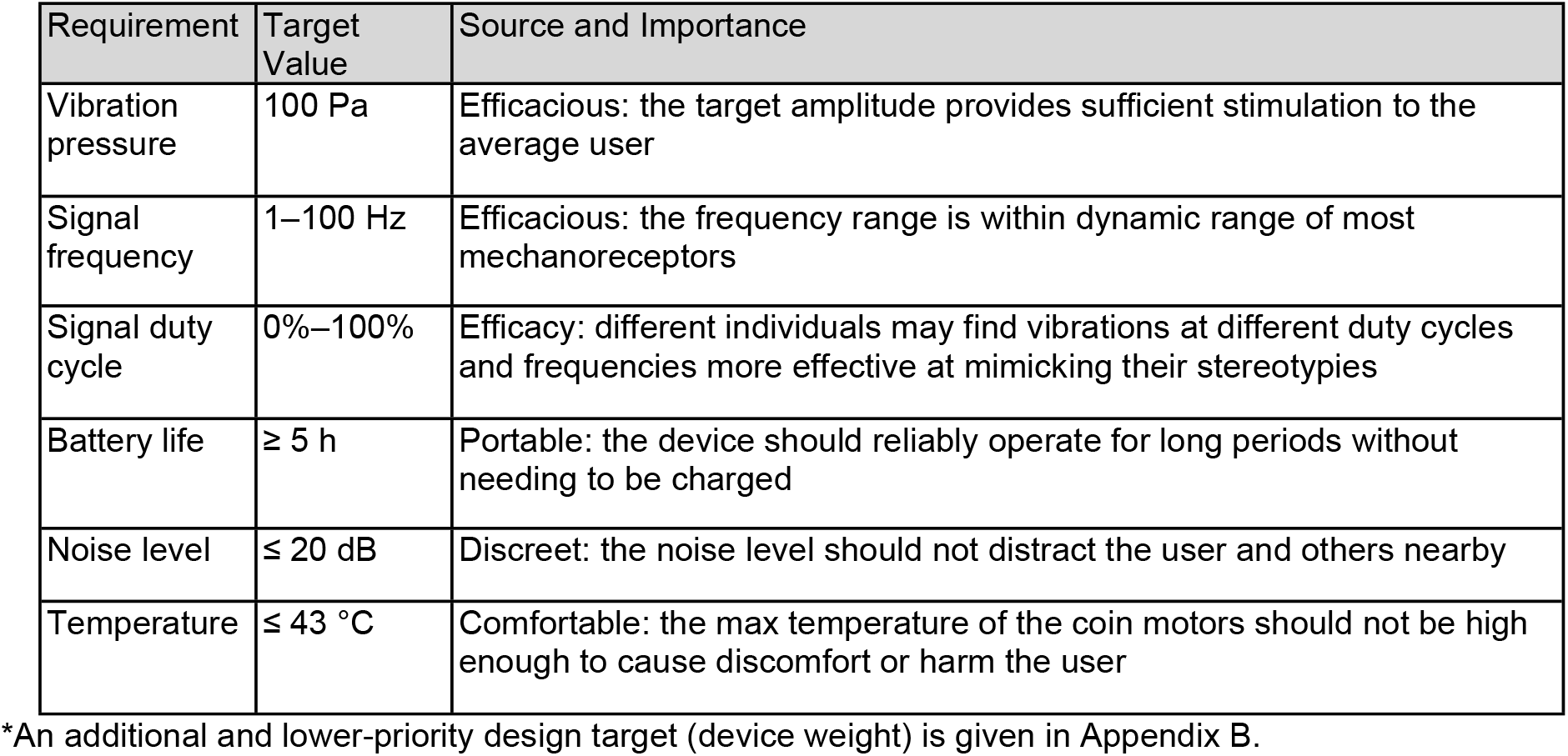
Design Targets*.

### Design Solution

#### Overview

Our design solution is a combination of two existing methodologies that have reduced stereotypy intensity and frequency: compressive deep pressure vests and vibration therapy. Our solution is a vibrating compressive vest with 18 small vibration motors distributed along the back, chest, and shoulders to stimulate a large surface area (see Fig. 1). Vibration frequency and duty cycle are adjustable in real time through a smartphone application to meet the user’s preferences and immediate needs.

**Figure 1.**
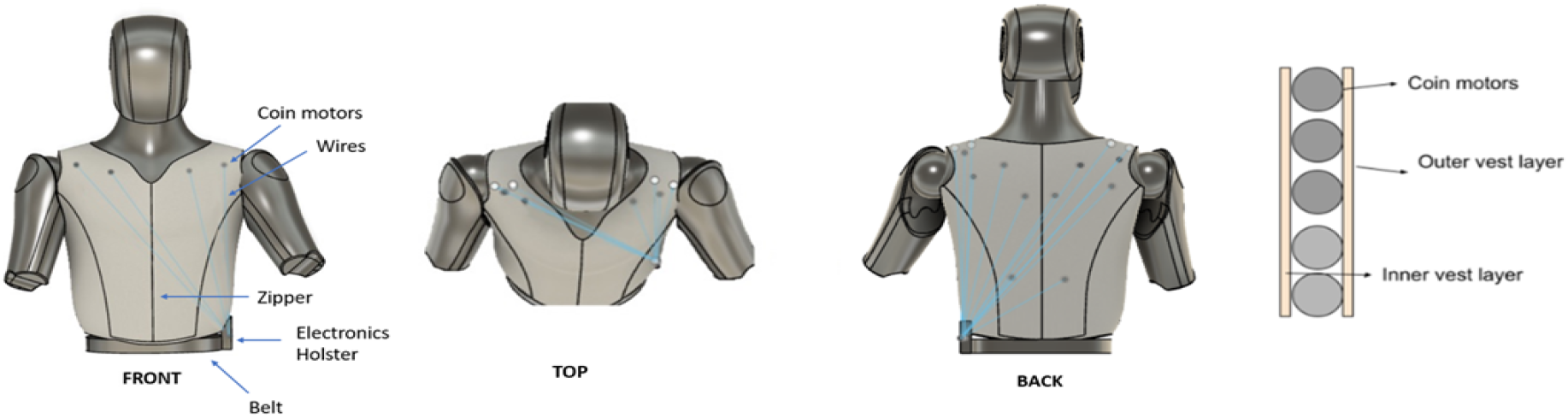
Front, top, and back view of the vibration vest design with coin motor placements (gray and white circles), wires (blue), belt, and holster containing the Arduino microcontroller, batteries, and Bluetooth module. Also displayed on the right is a graphic of the coin motors between the inner and outer layers of the compression vest.

A zipper allows the vest to be easily put on and taken off. In addition to vibration, the vest also uses elastic compressive material to provide snug compressive pressure to the user’s torso. The vest is thin enough for the user to wear under their normal clothing and has minimal noise levels, making it a discreet solution. An electronics holster houses the Arduino Uno microcontroller, batteries, and Bluetooth module.

### Detailed Description

#### Hardware

The foundation of the vest is a compression vest made of a stretchy, breathable material with a zipper through the middle that ensures ease of wear. The vest has 18 embedded coin vibration motors: upper chest (4), shoulders (4), lumbar (2), and trapezius (8). The number of motors in our prototype was limited by budgetary and supply chain constraints and technical limitations of the microcontroller (number of pins). We hypothesized that areas with moderate muscle density would be more sensitive to stimulation and requested motors to populate these regions. Motors are spaced approximately 1–2 inches apart to maximize surface area stimulated while also remaining in the designated areas. The coin motors and wires are attached to the vest using hook-and-loop fasteners and covered with a glued-on cloth layer so that neither is in direct contact with skin (see Fig. 1 rightmost image).

Wires and additional components are housed in a package disguised as a Leatherman-like holster connected to the bottom of the vest and attached to the user’s belt. The holster is a 47 × 77 × 122 mm, 3-D-printed polylactic acid box with a loop for attachment to a belt. The sliding lid of the box has a rectangular notch to accommodate for wire routing from the vest and a sliding switch. The box contains bulkier components that do not fit well on the vest, such as the batteries, Bluetooth module, microcontroller, sliding switch, and first nine transistors of each Darlington pair.

#### Electronics

There are 18 coin vibration motors used in this device. These 3 V DC motors measuring 8 mm in diameter are placed throughout the vest. They are controlled by the Arduino Uno microcontroller and 18 NPN transistors arranged as Darlington pairs. The microcontroller is connected to an HC-05 Bluetooth module, which receives information from a smartphone application to allow the user to control the motor groups, adjusting the duty cycle and vibrational frequency of pairs of coin motors. A circuit diagram of the prototype is given in Appendix D. The device is powered by two nickel metal hydride batteries with a combined capacity of 4,000 mAh with voltage outputs of 4.8 V. These batteries can power the device with all coin motors at full power for approximately 3.95 hours (calculation is given in Appendix E).

#### Software

The prototype is controlled by software on an Arduino Uno microcontroller (full code is given in Appendix F). The Arduino receives messages from a mobile phone application via an HC-05 Bluetooth module and controls the motor groups accordingly. After initialization, the Arduino enters the main loop (main code flowchart is given in the upper image of Appendix F), wherein it repeatedly checks whether a Bluetooth message has been received. If so, the Arduino parses the message and adjusts the on/off state, frequency, and duty cycle of each motor group accordingly. The frequency ranges from 1 to 100 Hz (inclusive) at 1 Hz intervals, and duty cycle ranges from 0% to 100% (inclusive) at 1% intervals.

The frequency and duty cycle of the motor groups are controlled with a 1 kHz interrupt service routine (ISR flowchart is given in the lower image of Appendix F). Every time the interrupt is triggered, the Arduino checks whether any of the motor groups need to be turned on or off. It does this by keeping track of a list of times (in ms) until each group needs to be toggled. When any time reaches zero, the Arduino will toggle the associated motor group and recalculate the time depending on that group’s frequency and duty cycle.

#### Phone Application

The user interface is an Android phone application with a single scrolling screen (see Fig. 2). It has a button at the top where the user can select the HC-05 Bluetooth module to connect to. It has a visual representation of the locations of the motor groups on the chest, shoulders, and back that users can click to turn on and off. Users can click and hold on one of the buttons to change the frequency and duty cycle of that single motor group or click the button “Change All Motors Vibration Speed” to control the frequency and duty cycle of all the motors at once.

**Figure 2.**
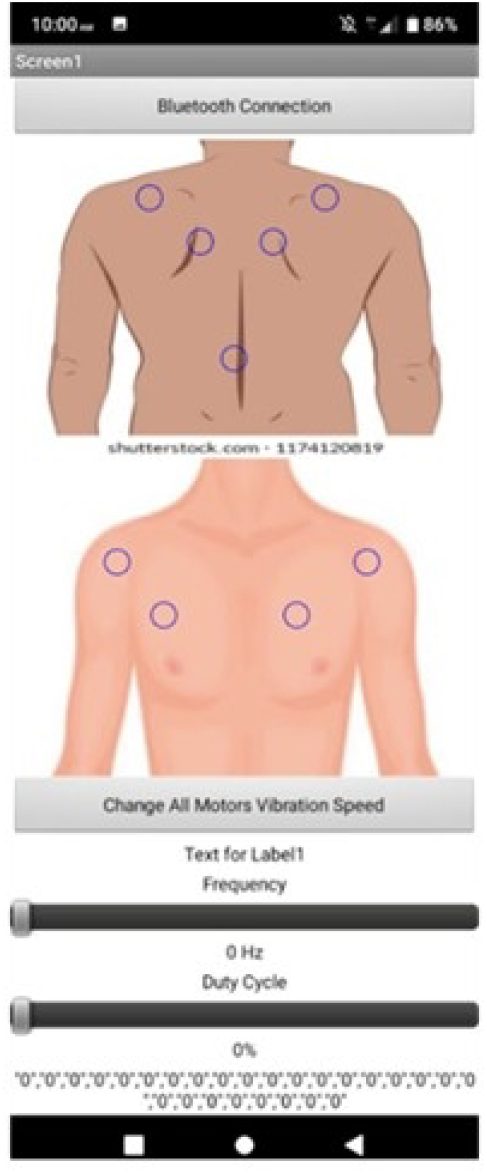
Smartphone application screen.

Once the motor group(s) are selected, the user can scroll down to the bottom of the screen to modify frequency and duty cycle using two sliders. These sliders allow for adjustment between 1 and 100 Hz for frequency and 0% to 100% for duty cycle and display the exact values of the sliders via labels that are constantly updated below them. Another label above the frequency slider indicates which motor region’s attributes are currently being modified.

Any time the motor groups, frequency, or duty cycle are changed, the application sends new user input data to the Arduino via the HC-05 Bluetooth module to apply the changes in real time so the user can quickly determine what settings they prefer. The full code is given in Appendix F.

#### Use

The vest is designed to be worn discreetly under regular clothing. The user should use the zipper to enclose the front of the vest. The package containing the electrical components such as the battery and microcontroller should be looped through the user’s belt. An application to control the vest must be installed on an Android smartphone, and the vest must be paired to the smartphone via Bluetooth. The smartphone can then act as a control to activate and alter vest vibration. This device should only be used with user consent and is not intended to be used continuously for lengthy periods. It should ideally be used when the individual would benefit from rhythmic sensory input that they would typically get from performing stereotyped movements.

### Evaluation

#### Prototype

The purpose of our final prototype iteration was a proof-of-concept prototype to test the software and circuit of our vest design. More specifically, the prototype would validate whether the frequency and duty cycle of the vibration motors can be adjusted accurately (< 5% error) with our software and circuit. It consists of the following components: Arduino Uno, Bluetooth module, coin vibration motor (×18), bipolar junction transistor (×18), compression vest, 4.8 V nickel metal hydride battery (×2), and a 3-D-printed electronics container.

The 18 coin vibration motors are split into nine groups that can be controlled independently via a mobile app. Whenever the user changes a parameter in the app, a Bluetooth message is sent to the Arduino, which updates the motor group specifications. Each group is controlled by an Arduino data pin signal and two NPN transistors arranged in Darlington pairs. The frequency and duty cycle timing is implemented via an interrupt-based timing mechanism.

### Performance and Testing Results

#### Vibration Pressure

The vibration intensity must be strong enough to penetrate muscle tissue and provide sensory stimulation to deeper nerves. We measured the peak pressure of the coin motors using a force-sensitive resistor (FSR). First, a baseline pressure was applied by pressing the FSR against a coin motor. Then, the motor was turned on at 1 Hz frequency and 50% duty cycle. This resulted in a square-wave pressure signal from which the baseline pressure was subtracted. The resulting peak pressure was 0.02 lbf/in2, or approximately 138 Pa, meeting our requirement of at least 100 Pa. We selected this value because we speculate the vibration pressure amplitude from smartphones to be appropriate for stimulation, which is typically approximately 100 Pa [33]. See Appendix E for a graph of the resulting data and for the Arduino code for the FSR.

#### Signal Frequency

Having a variable vibration frequency set by the user is important for mimicking the neurological feedback of stereotypies. Thus, it is important that the motors have an accurate frequency response according to the settings input by the user. The target range of 1–100 Hz was extrapolated from previous studies on vibration therapy and gives the user a range of different stimulation [25,26]. To test the accuracy, the prototype was set to various frequencies while at 50% duty cycle, and the motor control pin on the Arduino was sampled over a period of 3 seconds. The frequency of the signal was calculated in MATLAB (MathWorks Inc., Natick, MA, USA) using fast Fourier transforms (MATLAB code given in Appendix E), with the results shown in Table E1 of Appendix E. The percent error never exceeded 1%, indicating an accurate frequency response. However, the error seems to increase at higher frequency settings.

#### Signal Duty Cycle

A variable, user-defined duty cycle is also important in mimicking the neurological benefits of stereotypies and optimizing comfort. Therefore, the motors should have an accurate response to the changes in duty cycle. To test the accuracy, the prototype was set to various duty cycles at 1 Hz frequency, and the motor control pin on the Arduino was sampled over a period of 3 seconds. The duty cycle was estimated in Excel by calculating the area under the signal curve, with the results shown in

Table E2 of Appendix E. The percent error never exceeded 2%, indicating an accurate duty cycle response. However, it is likely that the duty cycle error will increase if tested at higher frequencies.

#### Battery Life

A long battery life is essential for a worn device designed for daily use. We estimated the battery life based on the specifications of our components. The result (given in Appendix E) was a battery life of 3.95 h, which does not meet our threshold of at least 5 h. However, this calculation assumes that all 18 motors are set at 100% duty cycle, which we do not expect in daily use. Thus, the practical battery life of our device will likely be much longer than the calculated value. We were not able to physically test the battery due to lack of battery-charging equipment.

#### Noise Level

A low noise level should be maintained to minimize public disruption and protect user privacy. We quantified the noise using an online decibel meter running on a laptop near the device, using the laptop’s microphone as input. We established a baseline noise level by letting the decibel meter run with the motors off. Then, we turned the device on at 1 Hz and 50% duty cycle and took the maximum difference between the new measurement and the baseline value. We made two measurements: one with the motors uncovered, and one with pressure being applied to the motors (which better mimics how the device will perform during use). The maximum noise level of the first case was 10 dB, and the maximum noise level of the second case was 5 dB. They both met our limiting value of 20 dB, which is equivalent to the noise level of someone whispering from 5 feet away.

#### Temperature

A noticeable difference in temperature occurs because the coin motors run continuously. To ensure the motors do not reach an unsafe temperature, we used an infrared camera to track the temperature of one coin motor running at 100% duty cycle over 5 minutes. A photo was taken of the motor every 30 seconds (given in Appendix E). The coin motor did not exceed the target temperature of 43 °C, determined by standard IEC 60601-1 as basic safety and essential performance of medical electrical equipment that has continuous contact with the user for over 10 minutes.

### FDA, Regulatory, and Ethical Considerations

FDA approval is not required because our device is classified as a class I device: it is a noninvasive device with low risk, is not intended to support life or prevent impairment to human health, and does not pose an unreasonable risk of injury. Our sponsor may decide how to best use the prototype for future clinical trials, including creating a grading system to quantify how users feel while wearing the device and whether this has any effect on stereotypy frequency/intensity.

For ethical considerations, we met with Sal Silinonte, an Autism Speaks Advocacy Ambassador and board of directors member for the Autism Society of Texas. Mr. Silinonte emphasized that the device should have strict ethical guidelines with a clear, well-defined mission statement. He emphasized that users should consent to using the device and that it should not be used as a restraint. He also emphasized that users should have full control over the device so they can self-direct and self-regulate according to their needs.

## Conclusion

We describe a prototype for a vest to be worn discreetly under clothing that provides rhythmic vibrotactile stimulation and have identified areas for improvement. One of the major limitations with our design is the wiring. Because the vest is a prototype, wiring is highly unoptimized. A wire is required for each communication channel, ground, and power line, which results in a bundle of 11 wires from the holster to the vest. This makes the vest bumpy and uncomfortable. The device has no external port for charging, so the user must remove the batteries from the device and charge them manually. Loose tolerances designed into the box to accommodate for variations in component selection resulted in a very visible, large box, which will likely make the vest more difficult to handle and wear.

By fabricating a custom flexible printed circuit board and using surface-mounted components, the device can be more comfortable, easier to fabricate, and less complex. Serial or I2C communication would further reduce the number of wires to the vest. User experience could be improved by making the vest detachable from the holster and integrating the charging module, allowing the user to charge the vest without removing the batteries. The user interface could be improved by having an option to preset frequency, duty cycle, and motor selection so that the device is easier to use.

Future improvements can increase the number of features. Motors may be encapsulated in padding to distribute vibratory force, dissipate heat, and improve comfort. Pockets for additional weights and a network of elastic webbing and velcro can provide adjustable deep pressure. A feature to synchronize vibration patterns with music could be added as a novelty or for therapeutic benefit. Finally, we foresee future closed loop versions coupled with EEG or other technologies to activate the device and adjust frequency and pressure according to physiological variables such as brain rhythms. This would be especialy useful to achieve nonintuitive patterns or to help people who are not able to fine-tune the stimulation parameters due to challenges in motor control or intellectual disability.

## Data Availability

All data produced in the present work are contained in the manuscript.

## Acknowledgments

Thank you to Dr. James Tunnell and Rachel Somavarapu (The University of Texas at Austin) for guiding and encouraging us throughout the design process. We thank Christina Roth for editorial and formatting assistance. Finally, we express our sincere appreciation to Sal Silinonte (Autism Society of Texas) for his input on the ethical and nontechnical considerations of our device.

## Appendix A

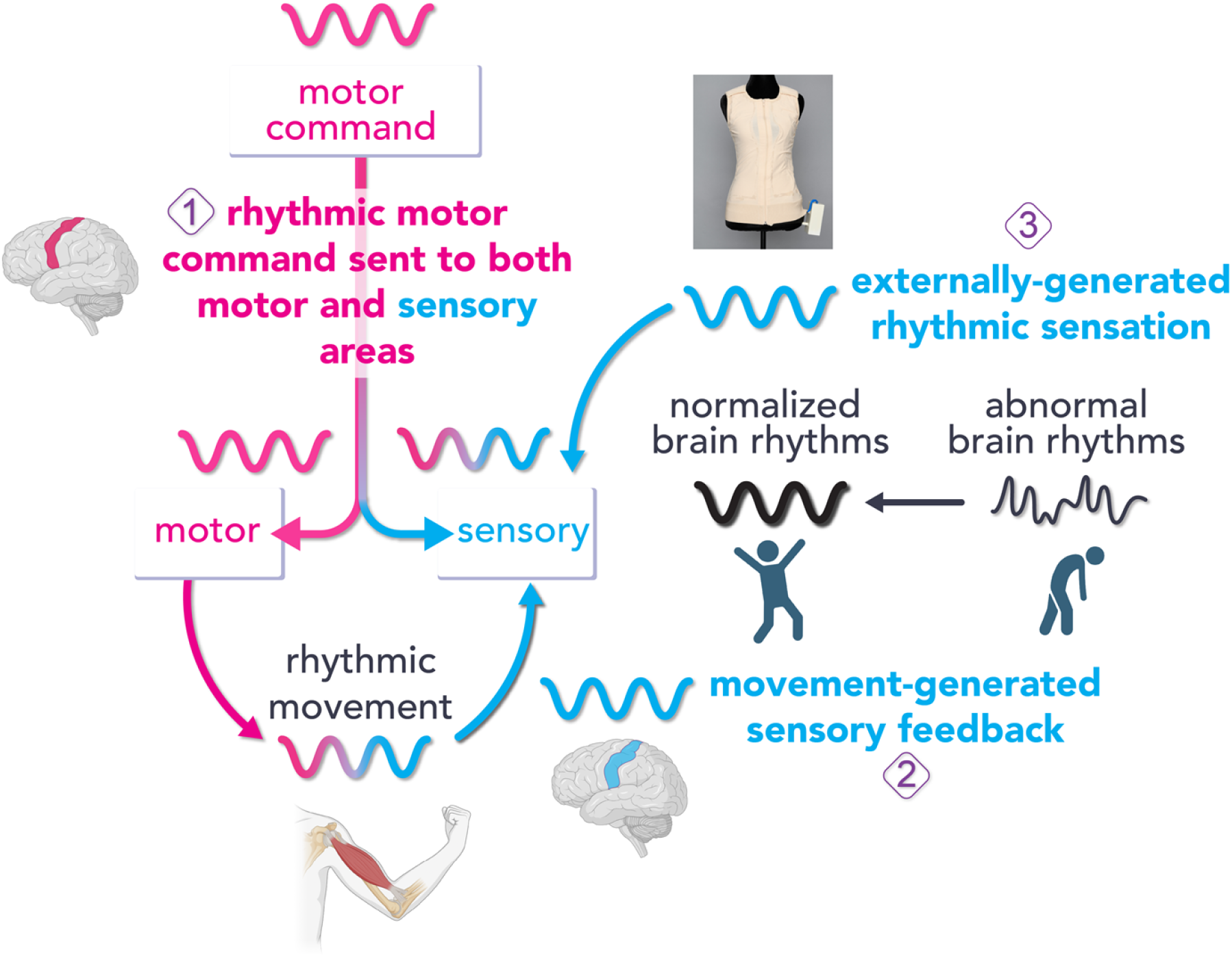

**The hypothesized signal pathway for the therapeutic benefit of rhythmic sensory inputs**. Adapted from McCarty & Brumback 2021. We hypothesize that rhythmic sensory signals can regulate brain rhythms by 3 mechanisms: **1)** Efference copy allows the brain to interpret sensory signals as self-generated vs. non-self-generated. To signal that a movement is self-generated, motor command signals are simultaneously sent to both the motor system and the sensory system. The copy of the motor command provides the sensory system a “heads up” that the movement was generated from within. Thus, **efference copy** provides rhythmic input to the sensory system. **2)** The **rhythmic movements** produce rhythmic sensory feedback. 3) An **external device** provides rhythmic sensory input. This model posits that rhythmic sensory input may serve as a therapeutic tool to normalize brain rhythms.

## Appendix B

Device Weight Requirement

**Appendix B: Device Weight Requirement**
| Requirement | Target Value | Source and Importance |
| --- | --- | --- |
| Device weight | $\leq 1$ kg | Portable: the device should be light enough to comfortably wear |

## Device Weight Testing

The device weight was measured by placing all components (the vest with all electronics attached and the 3-D-printed holster) into a plastic bag and weighing it on a spring scale. The result was 750 g, which meets our limiting value of less than 1 kg. People with autism sometimes prefer heavier clothing, so the weight could be an advantage.

## Appendix C

Electronic holster

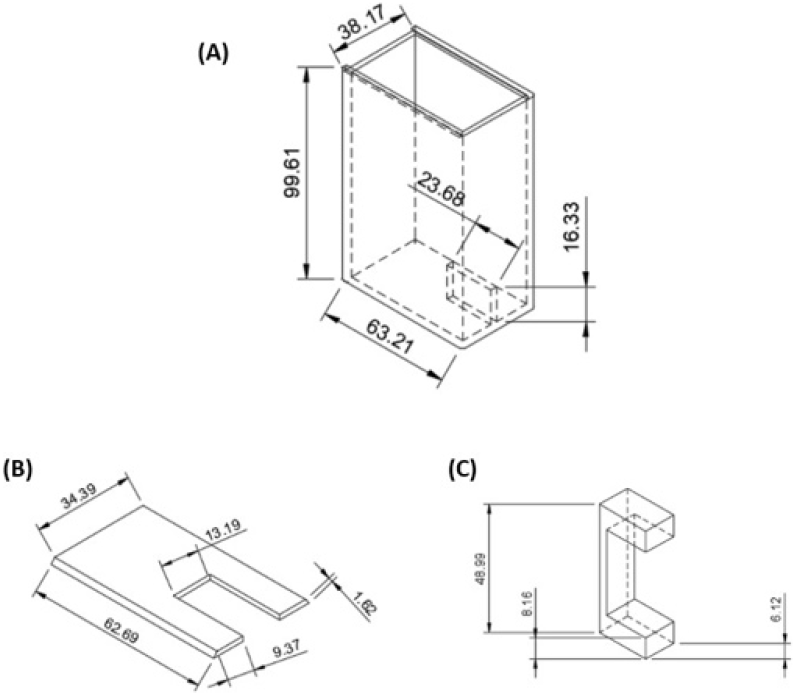

**CAD drawing of electronics holster components**. (a) Box with divider between Arduino and batteries, (b) lid that slides onto box with a hole for wires and power switch, and (c) belt loop that is superglued onto the box to make the holster wearable.

## Appendix D

Circuit diagram

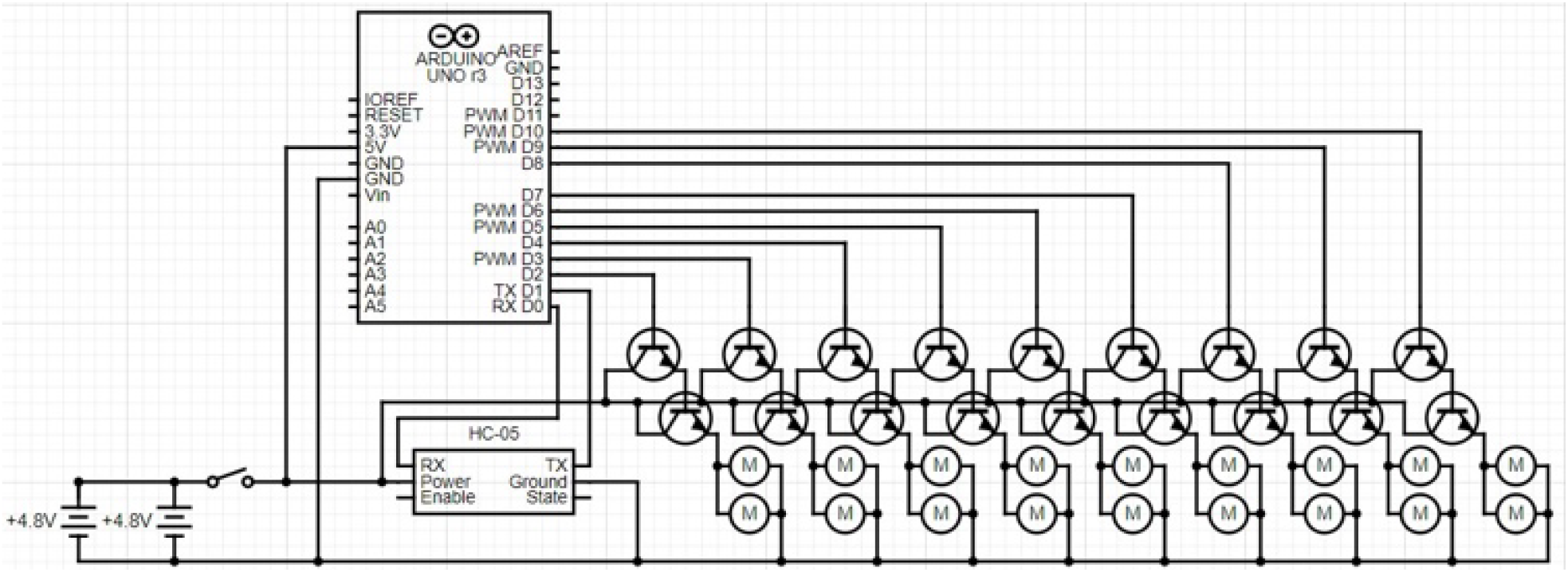

**Prototype circuit diagram**.

## Appendix E

Performance and Testing

## Vibration Pressure

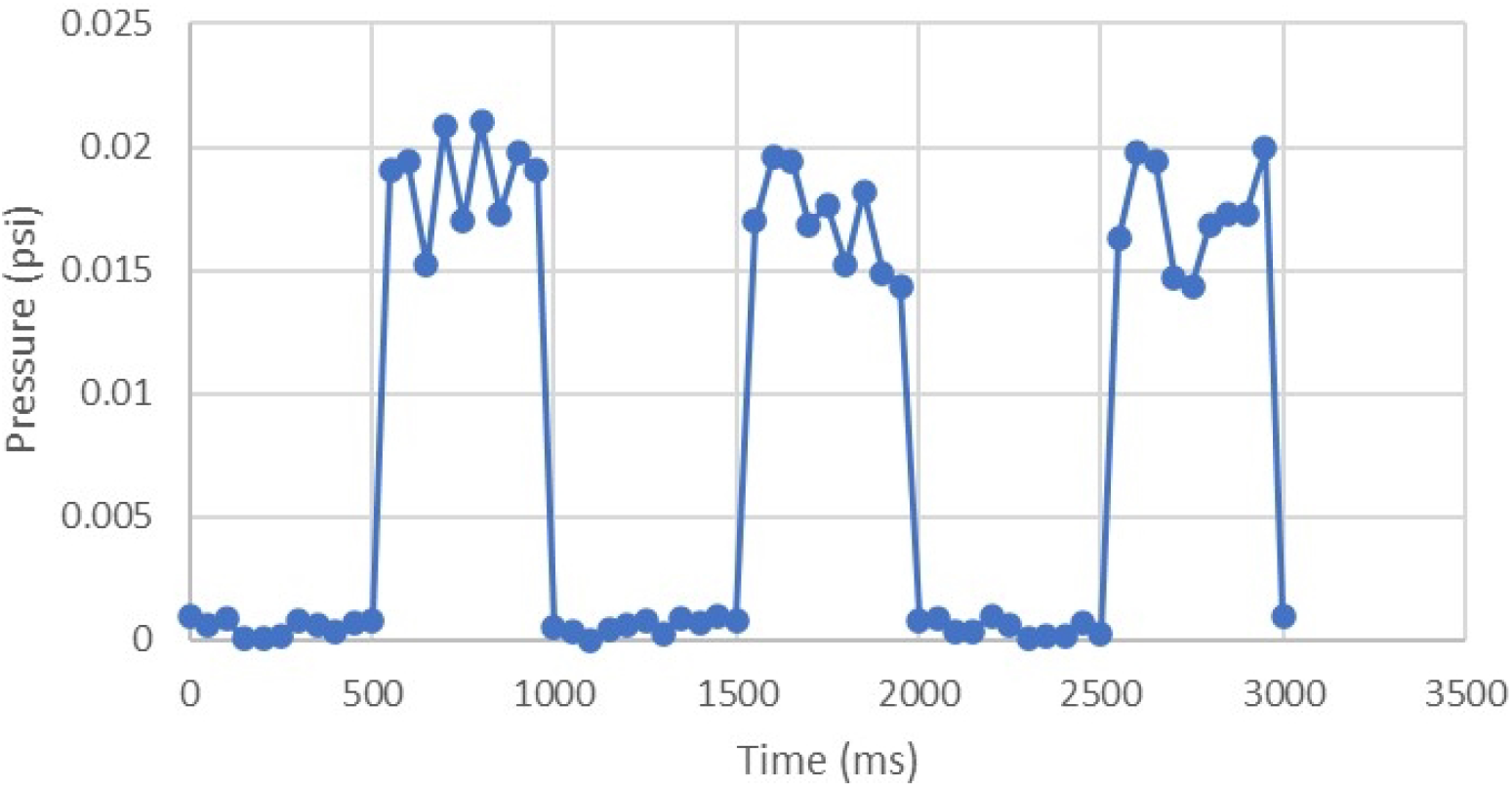

**Pressure testing for one coin motor vibrating at 1 Hz frequency and 50% duty cycle**.

## Signal Frequency

| Expected (Hz) | Actual (Hz) | Percent Error (%) |
| --- | --- | --- |
| 1 | 1.0013 | 0.130 |
| 2 | 2.0027 | 0.135 |
| 5 | 5.0067 | 0.134 |
| 10 | 10.013 | 0.130 |
| 25 | 25.033 | 0.132 |
| 50 | 49.733 | 0.534 |

**Frequency Accuracy Testing**. All frequency tests were performed at 50% duty cycle.

## Duty Cycle

| Expected (%) | Actual (%) | Percent Error (%) |
| --- | --- | --- |
| 5 | 4.940 | 1.20 |
| 25 | 24.87 | 0.52 |
| 50 | 50.15 | 0.30 |
| 75 | 75.08 | 0.11 |
| 95 | 94.93 | 0.07 |

**Duty Cycle Testing**. Results of duty cycle accuracy testing. All duty cycle tests were performed at 1 Hz frequency.

## Battery Life Calculation

(1a) *V* × *mAh*= *mWh*

*Battery Life*= *Power Storage Capacity*/*Power Draw*

## Coin Motors (3 V and 90 mA)

*Power Draw* = 18 *motors* × 3 *V* × 90 *mA*= 4. 86 *Wh*

## Battery Capacity (4000 mAh)

*Power Storage Capacity* = 4000 *mAh*× 4. 8 *V* = 19. 2 *Wh*

## Battery Life

*Battery Life*= 19. 2 *Wh*/4. 86 *Wh* = 3. 95 *hr*

## Temperature

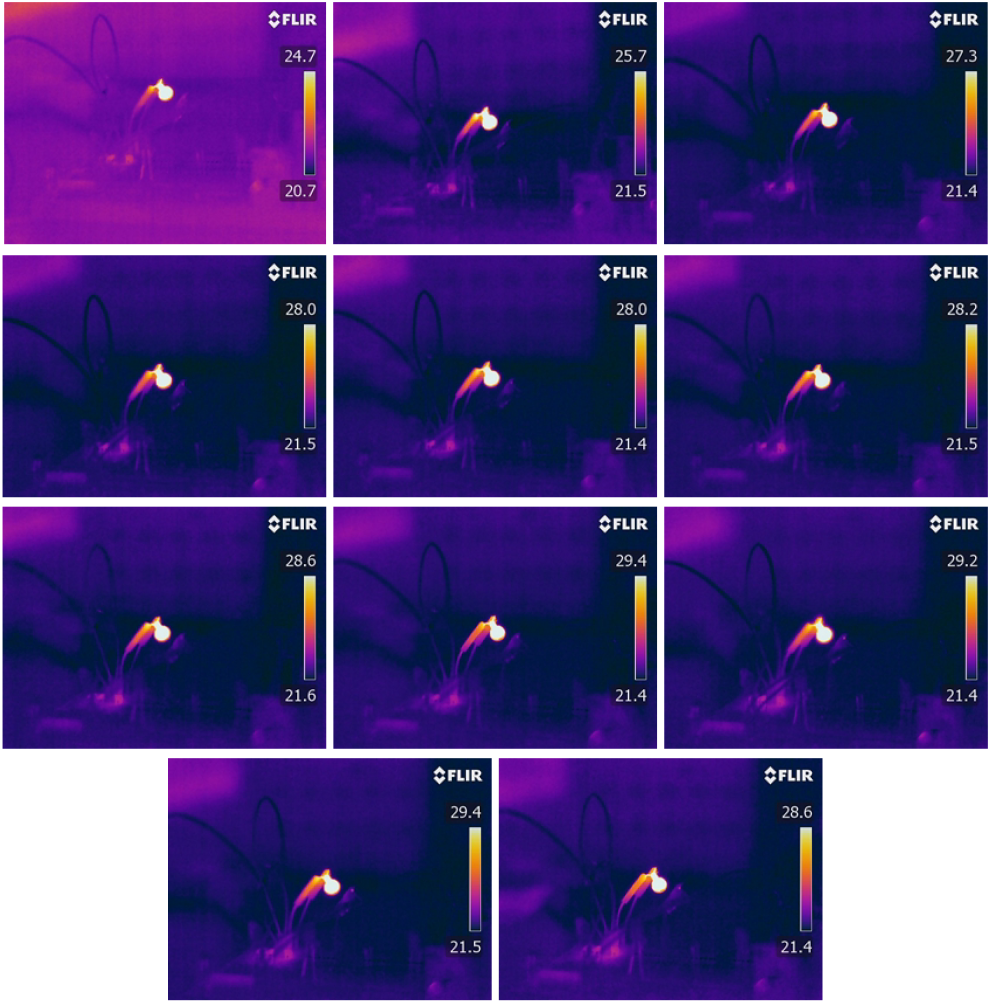

**Infrared images of one coin motor running at 100% duty cycle**. Pictures were taken every 30 seconds for 5 minutes (order: left to right, then top to bottom).

## MATLAB Frequency Estimate Code

~~~
%% Calculate Frequency of Signal
file = “Frequency and Duty Cycle Testing v3”; sheet = “freq50”;
% Read data from Excel sheet data = xlsread(file, sheet); time = data(:,1); val = data(:,2);
% Create new variables for padding out time2 = 0:time(end); val2 = zeros(1, length(time2));
% Fill in gaps from original data (emulate sampling at 1 kHz) ind = 1; for i = 1:length(time2)
if time(ind) ∼= time2(i) val2(i) = val2(i-1); else
val2(i) = val(ind); ind = ind + 1; end
end
% Perform fast fourier transform L = length(time2);
y = fft(val2);
p2 = abs(y/L); p1 = p2(1:L/2+1);
p1(2:end-1) = 2*p1(2:end-1);
f = 1000*(0:(L/2))/L;
plot(f, p1);
% Output frequency estimate [∼, index] = max(p1(1:end)); f(index)
~~~

## Pressure Sensor (FSR) Code

~~~
// Pressure Sensor (FSR) Code
// One end of FSR connected to +3.3V, other end connected to A0 with pull-down resistor
const int analogInPin = A0;
//const int analogOutPin = 9; const int R1 = 10050;
double R2;
int sensorValue = 0; int outputValue = 0; double voltage = 0; double pressure;
void setup() {Serial.begin(9600);
}
void loop() {
sensorValue = analogRead(analogInPin); outputValue = map(sensorValue, 0, 1023, 0, 255);
//analogWrite(analogOutPin, 255); voltage = 3.3*outputValue/255;
R2 = R1 * ((3.3/voltage) -1.0);
//Serial.println(R2);
pressure = 23949*pow(R2, -1.31); Serial.println(pressure); delay(100);
}
~~~

## Appendix F

Arduino code

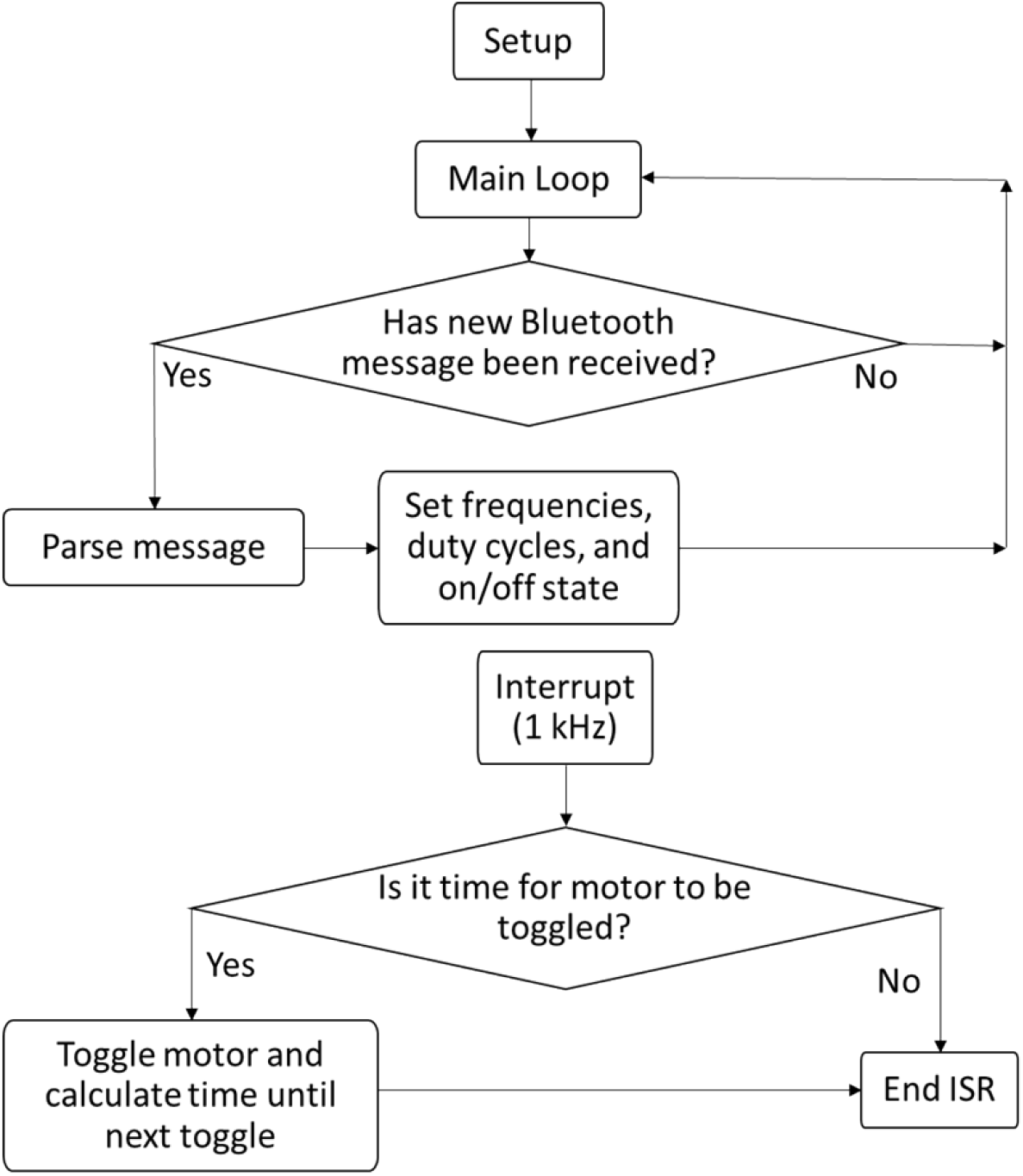

**Arduino Software Flowchart** with main device code (top) and ISR routine (bottom).

## Arduino Code

~~~
#define numRegions 2
#define minFreq 1
#define maxFreq 100
#define freqInc 1
#define minDuty 0
#define maxDuty 100
#define dutyInc 5
// Rotary encoder pin order
#define CLK 0
#define DT 1
#define SW 2
// Pins for each motor region
int regionPin[numRegions] = {2, 4};
// Pins for each rotary encoder module (CLK, DT, SW) int rotaryPin[numRegions][3] = {{3, 5, 6}, {9, 10, 11}};
// CLK states for each rotary encoder int CLKState[numRegions];
// SW states for each rotary encoder int SWState[numRegions];
// Mode of reach rotary encoder (true = frequency, false = duty cycle) bool rotaryState[numRegions] = {true, true};
// Initial settings for each motor region float freq[numRegions] = {1, 1}; int dutyCycle[numRegions] = {50, 50}; long timeLeft[numRegions];
// ******************************************************************************
// setup()
// Initializes pin modes and starting values
// ******************************************************************************
void setup() {
for (int i = 0; i < numRegions; i++) {pinMode(regionPin[i], OUTPUT);
pinMode(rotaryPin[i][CLK], INPUT); pinMode(rotaryPin[i][DT], INPUT); pinMode(rotaryPin[i][SW], INPUT_PULLUP);
CLKState[i] = digitalRead(rotaryPin[i][CLK]); SWState[i] = digitalRead(rotaryPin[i][SW]); timeLeft[i] = 10 * dutyCycle[i] / freq[i];
}
Serial.begin(9600); setupTimer1(1000);
}
// ******************************************************************************
// loop()
// Toggles motors on and off at a variable frequency and duty cycle
// ******************************************************************************
void loop() {
// Process rotary encoders and check if time to toggle motors for (int i = 0; i < numRegions; i++) {processRotary(i);
}
}
// ******************************************************************************
// processRotary()
// Reads rotary encoder and adjusts frequency or duty cycle accordingly
// Input: region ID
// ******************************************************************************
void processRotary(int region) {
int CLKval = digitalRead(rotaryPin[region][CLK]); int DTval = digitalRead(rotaryPin[region][DT]); int SWval = digitalRead(rotaryPin[region][SW]);
// If last and current state of CLK are different, then pulse occurred if (rotaryState[region]) {
// Frequency mode
if (CLKval != CLKState[region] ×× CLKState[region] == 1) {if (DTval == CLKval ×× freq[region] > minFreq)
freq[region] -= freqInc;
else if (freq[region] < maxFreq) freq[region] += freqInc;
}
} else {
// Duty cycle mode
if (CLKval != CLKState[region] ×× CLKState[region] == 1) {if (DTval == CLKval ×× dutyCycle[region] > minDuty)
dutyCycle[region] -= dutyInc;
else if (dutyCycle[region] < maxDuty) dutyCycle[region] += dutyInc;
}
}
CLKState[region] = CLKval;
// Button has been released
if (SWState[region] == LOW ×× SWval == HIGH) rotaryState[region] = !rotaryState[region];
SWState[region] = SWval;
}
// ******************************************************************************
// ISR(TIMER1_COMPA_vect)
// Toggles motors on and off
// ******************************************************************************
ISR(TIMER1_COMPA_vect) {
for (int i = 0; i < numRegions; i++) {
if (dutyCycle[i] == 0) {digitalWrite(regionPin[i], LOW); continue;
} else if (dutyCycle[i] == 100) {digitalWrite(regionPin[i], HIGH); continue;
}
timeLeft[i]--;
if (timeLeft[i] <= 0) {
if (digitalRead(regionPin[i])) {
timeLeft[i] = 10 * (100-dutyCycle[i]) / freq[i]; digitalWrite(regionPin[i], LOW);
} else {
timeLeft[i] = 10 * dutyCycle[i] / freq[i]; digitalWrite(regionPin[i], HIGH);
}
}
}
}
~~~

## Appendix G

**Smartphone Application Code**

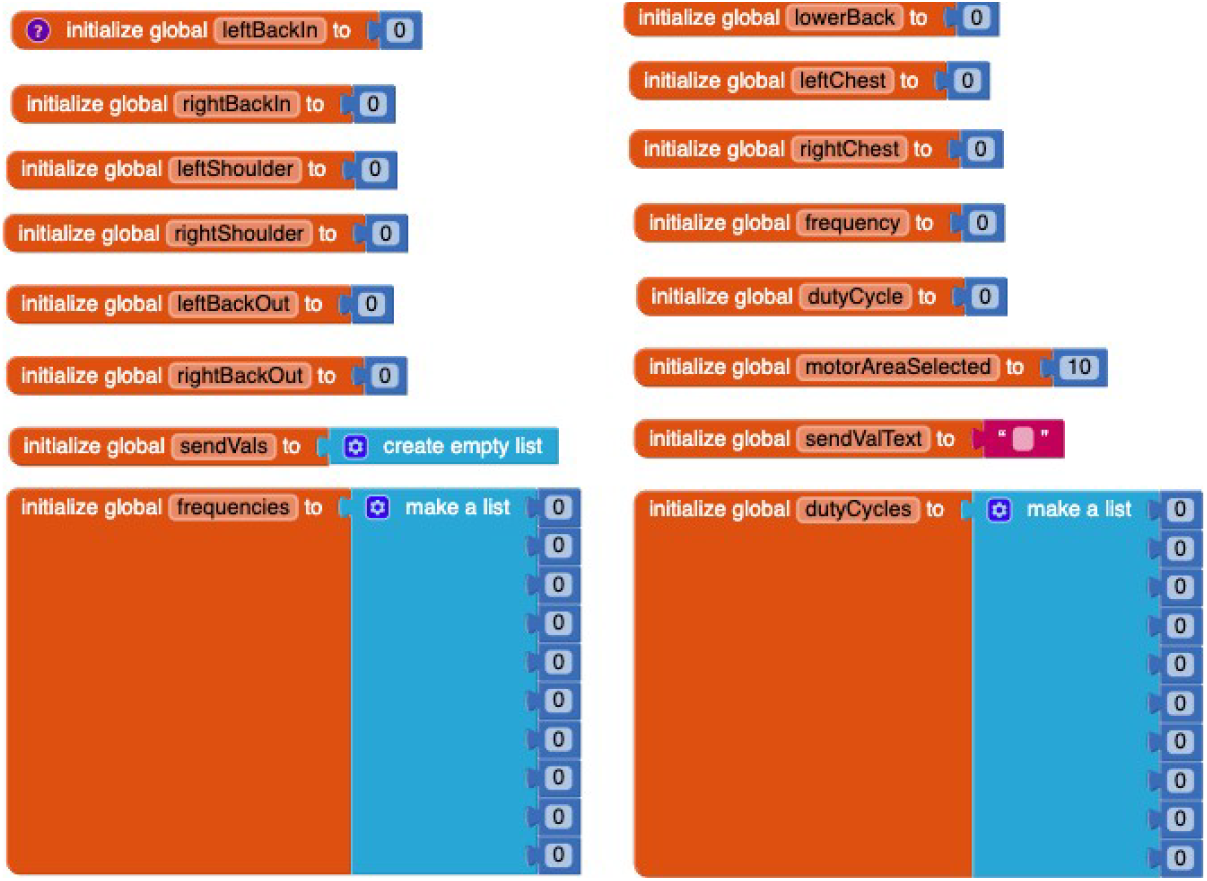

Initializes all variables needed for the user interface code.

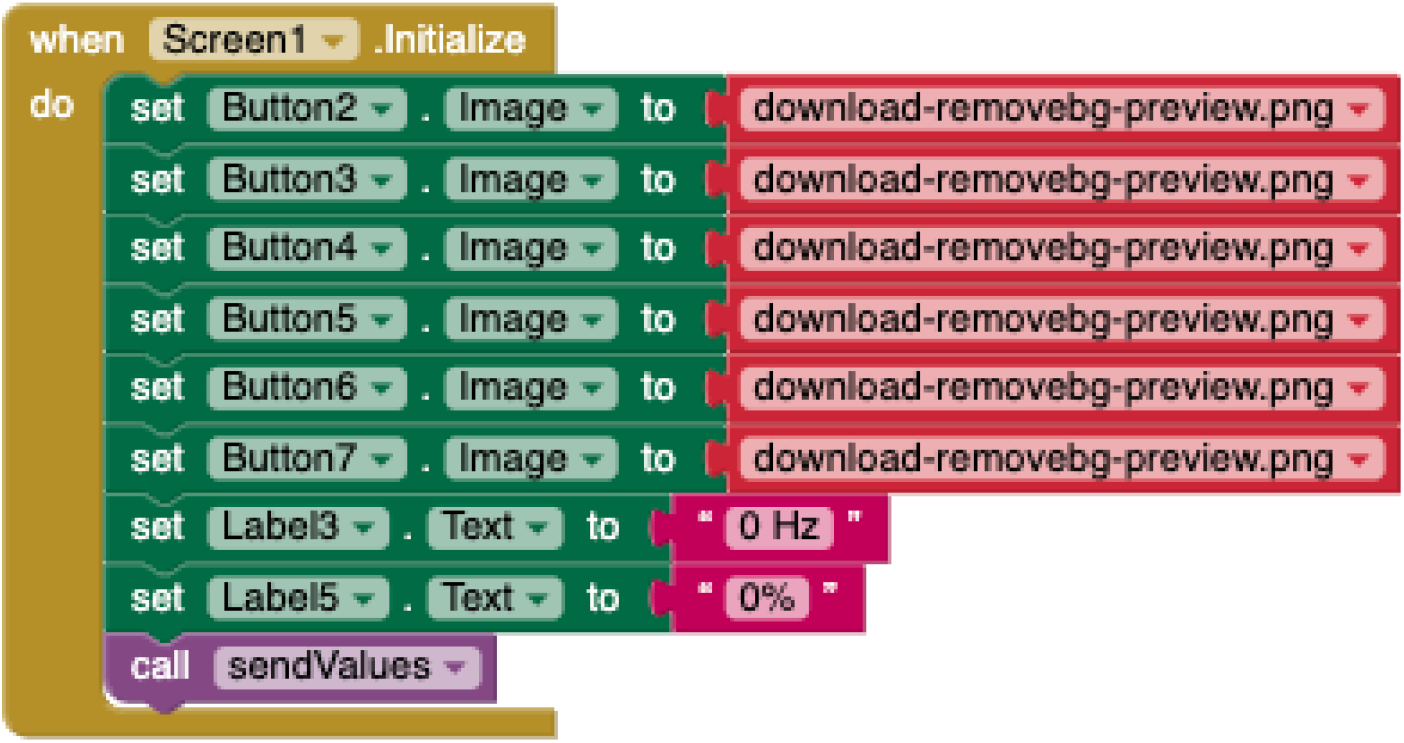

Initializes all buttons to show as not selected and initializes starting labels for frequency and duty cycle sliders.

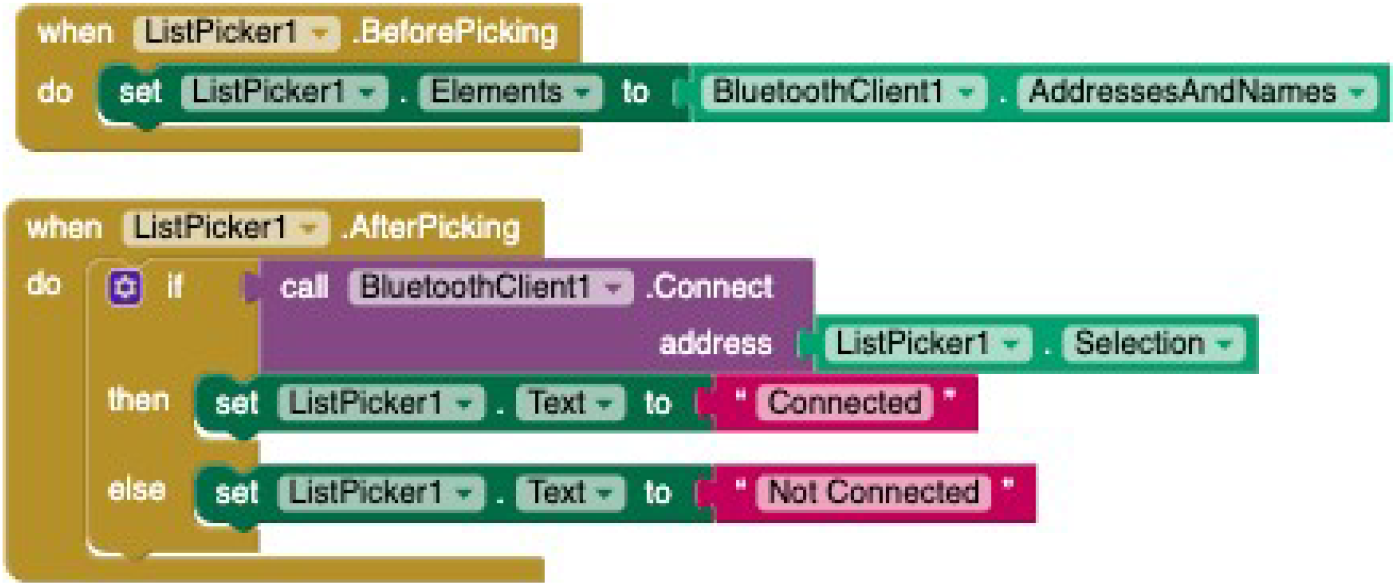

Allows user to set up Bluetooth connection to HC-05 Bluetooth module from phone application.

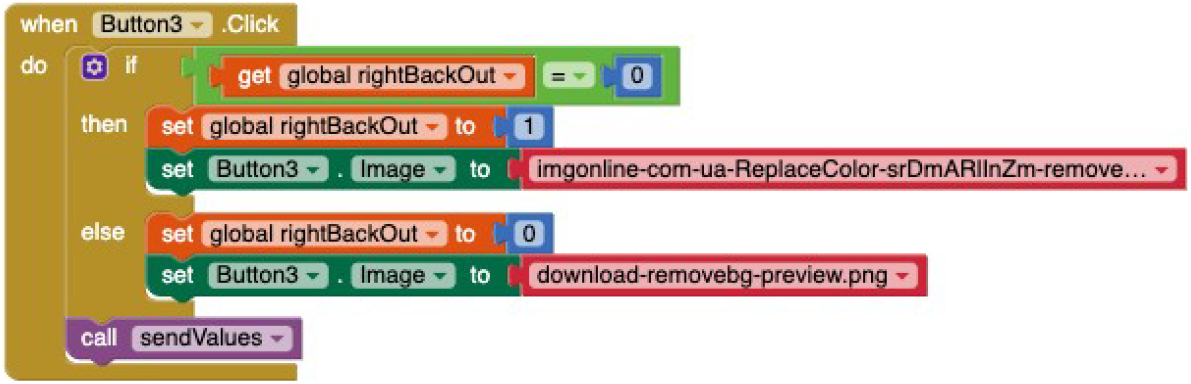

Allows user to click certain buttons to turn on certain motor areas, repeated for all motor area buttons.

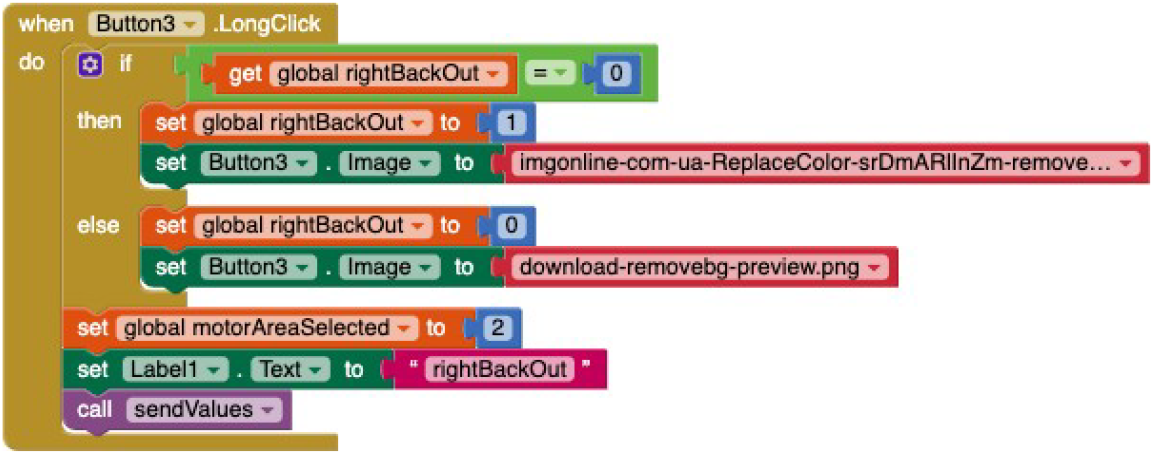

Allows user to click and hold to select a motor area and then change the frequency and duty cycle for only that motor area, repeated for all motor area buttons.

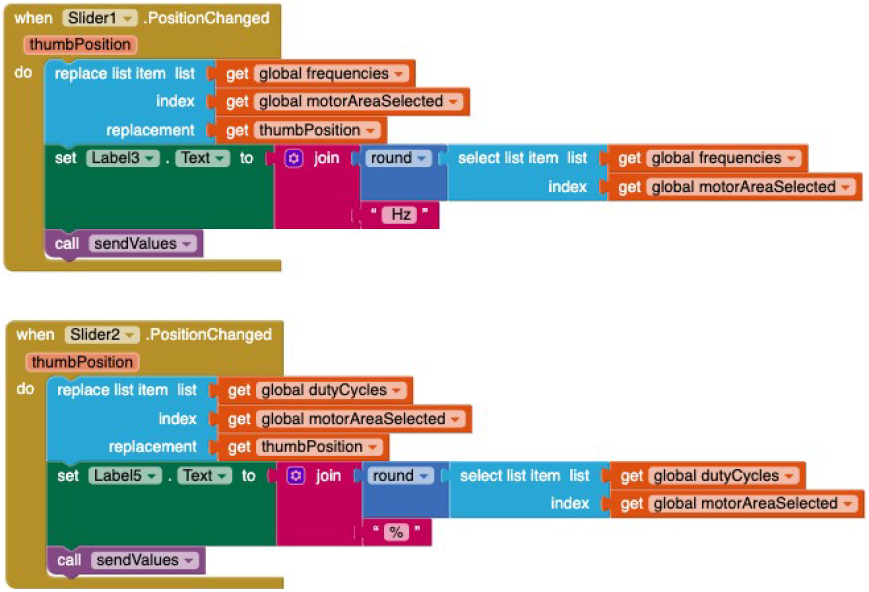

Allows user to modify frequency and duty cycle using sliders of a specific motor area or all motors based on what was previously selected.

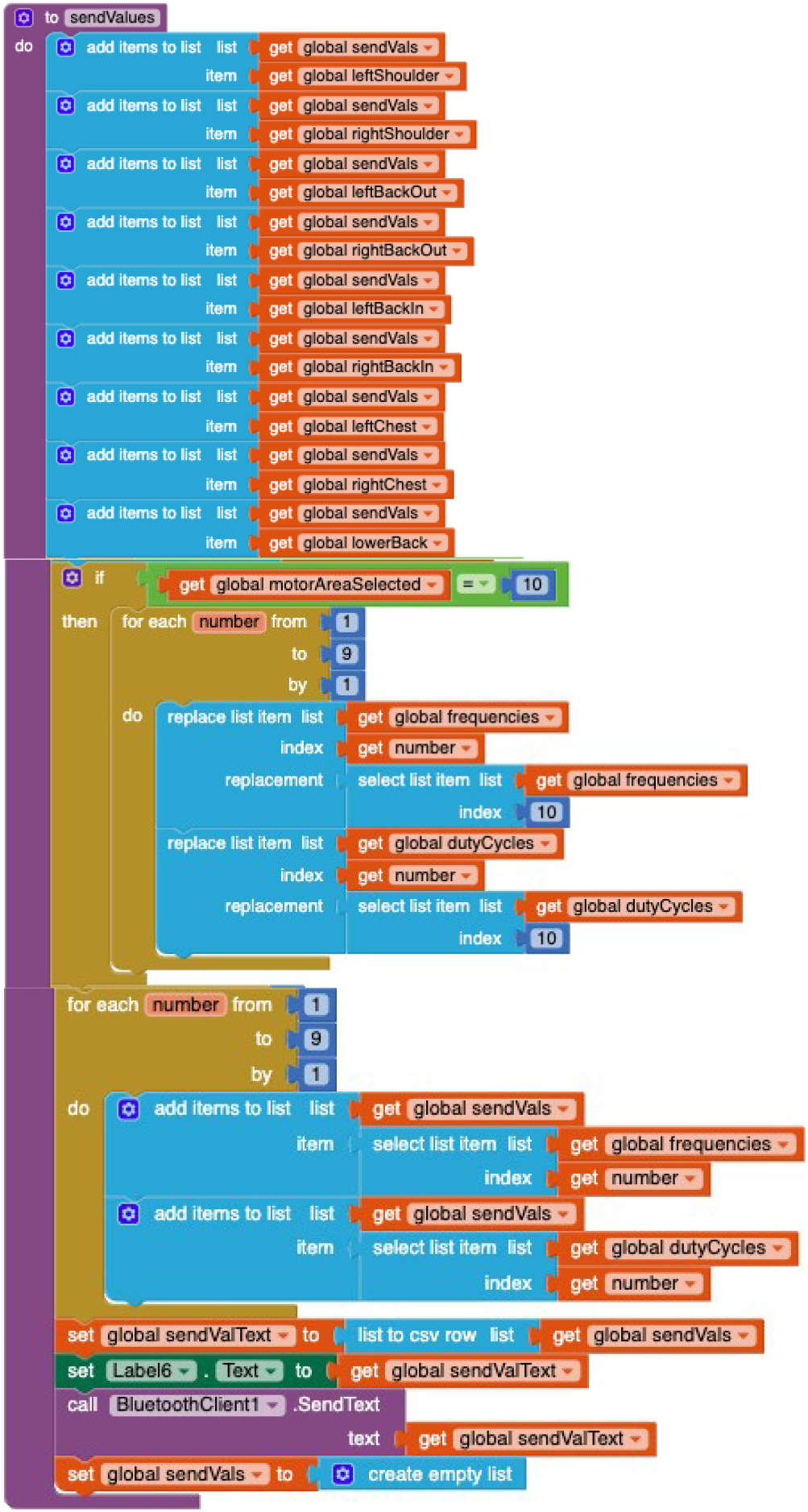

sendValues function is called every time a user modifies a parameter on the application and sends motor areas selected, frequencies, and duty cycles to the Arduino as a string, which are then parsed by Arduino code.

## Appendix H

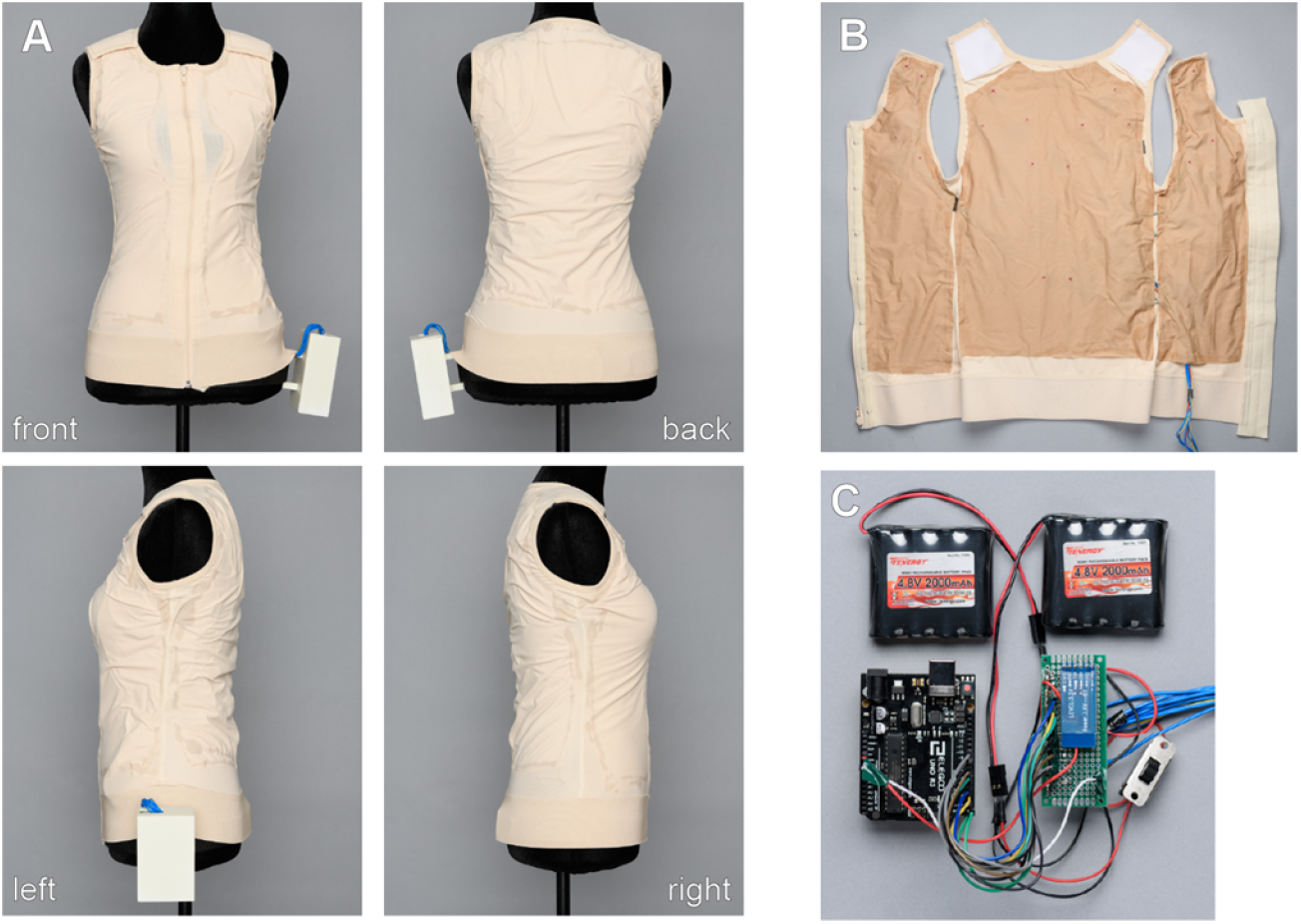

**Vest prototype**

**A**. Vest exterior. **B**. Vest interior. **C**. Batteries, Arduino Uno microcontroller, and Bluetooth module.

